# Kidney Protection During Surgery on the Thoracoabdominal Aorta: A Systematic Review

**DOI:** 10.1101/2024.06.25.24309413

**Authors:** James Thomas Bennett, Sarah Shirley, Patricia Murray, Bettina Wilm, Mark Field

**Affiliations:** Liverpool Heart and Chest Hospital NHS Trust, Liverpool, England; The University of Liverpool, Faculty of Health and Life Sciences, Liverpool, England

**Keywords:** Thoracoabdominal Surgery, Aorta, Kidney, Protection, Renal, Perfusion

## Abstract

**Background:** Acute kidney injury (AKI) is a common complication of surgery to repair the thoracoabdominal aorta, and is associated with risks of dialysis and early mortality. Renal ischaemia, initiated by clamping of the suprarenal aorta, is a major cause. Consequently, perfusion techniques are commonly used to sustain renal blood flow or facilitate hypothermic kidney preservation during surgery. This systematic review provides a comprehensive assessment of renal and mortality outcomes by perfusion techniques, to evaluate their ability to provide effective kidney protection.

**Methods & Results:** Searches of PubMed, Web of Science, ClinicalTrials.gov and ClinicalTrialsRegister.EU were conducted to identify relevant studies published from 1995 to 2024. Included studies were quality assessed, and data were extracted by perfusion techniques. Outcomes of the highest quality studies were used to synthesise a narrative discussion.

Forty-five studies were included in our analysis, featuring three extracorporeal strategies: Left heart bypass (LHB; n=24), cardiopulmonary bypass with deep hypothermic circulatory arrest (DHCA; n=19), and partial cardiopulmonary bypass (pCPB; n=12). Three categories of selective renal perfusion strategy were identified: Warm blood, cold blood and cold crystalloid. Our analysis identified operative mortality as 0-23.4% following LHB, 2.2-12.5% following DHCA and 0-42.1% following pCPB. The incidence of renal replacement therapy was 0-40.0% following LHB, 0-15.0% following DHCA and 0-22.2% following pCPB.

**Conclusions:** Strong evidence supports the use of distal aortic perfusion (DAP) with LHB or pCPB, to reduce the risks of dialysis and operative mortality associated with aortic cross clamping. Furthermore, when DAP cannot prevent kidney ischaemia, adjunctive perfusion of the renal arteries with cold histidine-tryptophan-ketoglutarate (HTK) can reduce the risk of AKI. However, no professional guidance on the management of HTK exists, and rates of AKI remain high despite its use. Selective renal perfusion with warm blood is identified as a risk factor for AKI and operative mortality. DHCA is associated with low rates of AKI, warranting further prospective investigation. Finally, intravascular haemolysis and myoglobinaemia are acknowledged as important risk factors that require urgent research to address the problem of AKI.

## Introduction

### Background

Surgical repair of the thoracoabdominal aorta (TAA) (figure 1) is a life-saving procedure for the treatment of aneurysms that present patients with high risks of rupture or visceral organ ischaemia^1, 2^. However, this complex undertaking can expose patients to several physiological challenges, including ischaemia-reperfusion injury, intravascular haemolysis, and acute inflammation^3–5^. Such factors can contribute to the development of post-operative organ dysfunction, and may manifest as disorders such as acute kidney injury (AKI), paraplegia, stroke or pulmonary dysfunction^6^. Reducing the incidence of such complications is essential to improve patient quality of life (QOL) following surgery^6^.

**Figure 1.**
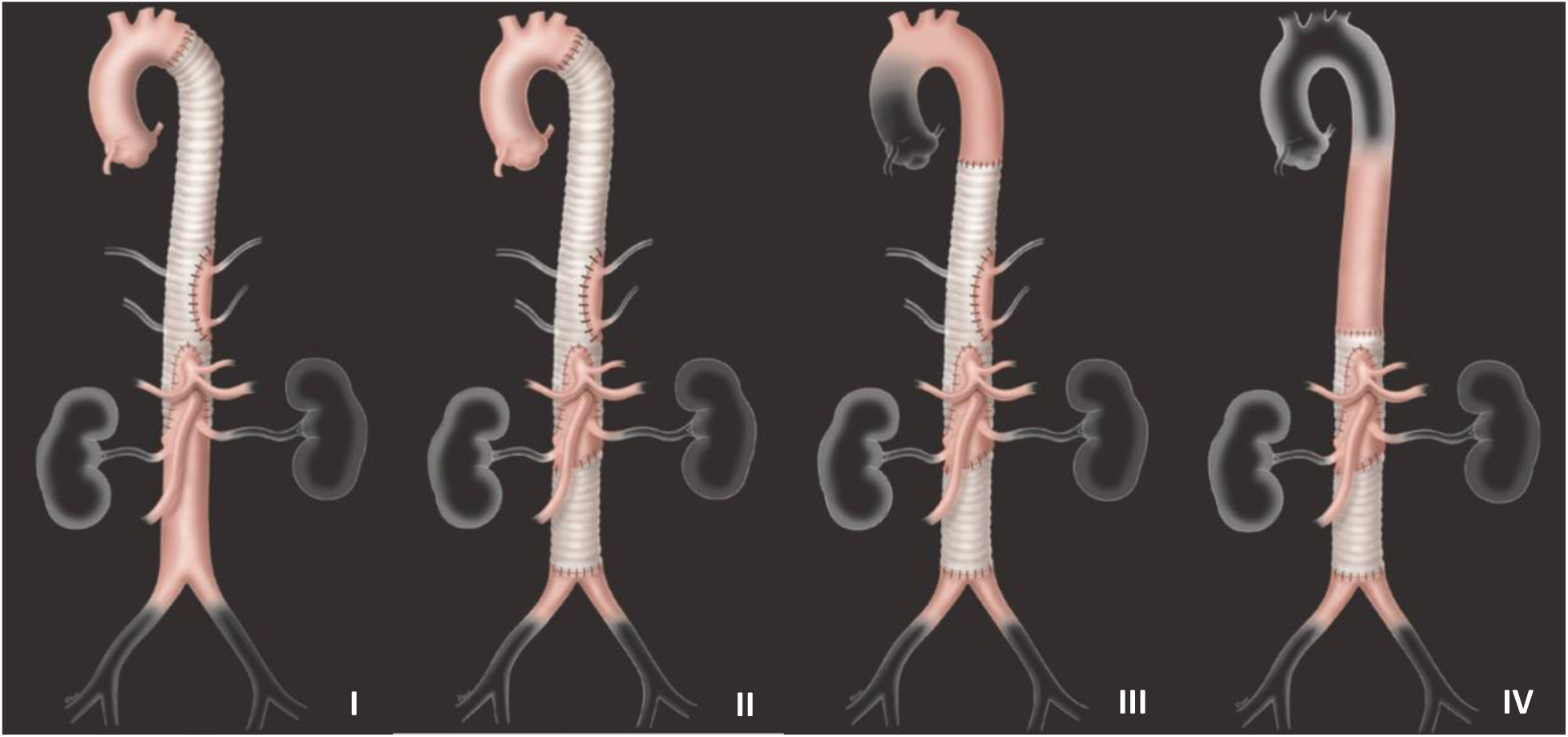
Crawford extents of the thoracoabdominal aorta.

Intra-operative organ protection strategies can be used to improve patient care^7^. For instance, the use of distal aortic perfusion (DAP), cerebrospinal fluid drainage and intra-operative neuromonitoring can ensure effective spinal cord protection, and reduce the risk of post-operative paraplegia^8–12^. In contrast, providing effective intra-operative renal protection has proven a more elusive ambition; AKI has been reported to affect the majority of patients who undergo TAA repair^13, 14^. As AKI is a significant predictor of early mortality^15^, establishing effective renal protection strategies is imperative to improve patient QOL and operative survival.

### Acute Kidney Injury

AKI denotes an abrupt decline in renal function, diagnosed by an elevated serum creatinine concentration and/or decreased urine output (table 1)^16, 17^. Several classification systems exist, including KDIGO, RIFLE and AKIN, to score the severity of AKI from mild to severe^18^. Severe AKI typically requires the use of renal replacement therapies (RRT), such as haemodialysis, and is associated with the greatest risk of operative mortality^15, 19–22^. However, even mild AKI can increase a patient’s risk of early death^18^. Furthermore, AKI can progress towards chronic kidney disease, with up to 7% of TAA patients requiring RRT following hospital discharge^22–25^. Understanding how AKI develops is essential to design targeted strategies for its prevention^26^.

**Table 1:** KDIGO AKI classification criteria for adults^17^.

| **Table 1: KDIGO AKI classification criteria for adults<sup>17</sup>** |  |  |
| --- | --- | --- |
| **Stage** | **Serum creatinine concentration** | **Urine output** |
| 1 | 1.5-1.9 times baseline<br>OR $\geq 0.3\text{mg/dl}$ ($\geq 26.5\text{mmol/l}$)<br>increase | $<0.5\text{ml/kg/h}$ for 6-12 hours |
| 2 | 2.0-2.9 times baseline | $<0.5\text{ml/kg/h}$ for $\geq 12$ hours |
| 3 | 3.0 times baseline<br>OR<br>$\geq 4.0\text{mg/dl}$ ($\geq 353.6\text{mmol/l}$)<br>OR<br>Initiation of renal replacement<br>therapy | $<0.3\text{ml/kg/h}$ for $\geq 24$ hours<br>OR<br>Anuria for $\geq 12$ hours |

The pathophysiology of AKI is complex and multifactorial^27^. However, extended renal ischaemia, instigated by clamping of the suprarenal aorta, is the primary cause in the context of TAA surgery^7, 26, 28^. Clamping may be necessary to facilitate aortic transection and repair, but causes an abrupt pause in renal blood flow that can materialise as acute tubular necrosis and localised inflammation^26^. Additionally, intravascular haemolysis, caused by shearing forces within extracorporeal circuits, can cause renal endothelial dysfunction and contribute to kidney hypoperfusion^4, 29, 30^.

Renal ischaemia generates hypoxic conditions that deplete intracellular ATP and inhibit essential processes in all segments of the nephron^31^. Proximal tubular epithelial cells, however, are notably vulnerable to hypoxic injury, due to their dependence on oxidative phosphorylation to sustain the energetic processes of tubular reabsorption and secretion^16, 27, 32, 33^. Severe or extended hypoxia can induce cell death through apoptosis and necrosis^13, 16, 31^. Furthermore, subsequent re-perfusion of the hypoxic kidney, on removal of the aortic cross-clamp, can amplify the mitochondrial production of reactive oxygen species and activate pro-inflammatory pathways^16, 34, 35^. These processes can culminate in cell damage and dysfunction, collectively known as ischaemia-reperfusion injury (IRI), and inhibit glomerular filtration rate^27, 31, 35^.

### Intra-operative Kidney Protection

Open repair of the TAA can be performed with the independent use of ‘simple aortic cross clamping’ (SACC), in which no systemic circulatory support strategies are employed. However, with SACC, the period of suprarenal clamping directly corresponds to the duration of renal ischaemia, and augments the risk of AKI^21, 36, 37^. Additionally, SACC is related to an increased risk of neurological injury^38, 39^. Expedient surgery is therefore required to minimise organ ischaemia, and prevent AKI.

More commonly, extracorporeal perfusion strategies are employed to attenuate renal IRI through oxygenation or hypothermia (figure 2). Strategies of left heart bypass (LHB) or partial cardiopulmonary bypass (pCPB) can provide DAP and supply the kidneys with normothermic, oxygen-rich blood during surgery^28, 37^. Both strategies are analogous in their ability to perfuse the distal aorta, and their reliance on sustained cardiac output and ventilation to perfuse arteries proximal to the aortic cross clamp. However, they differ in their respective cannulation sites and perfusion techniques: LHB involves the active shunting of oxygenated blood from the left side of the heart to the distal aorta; pCPB utilises blood drained from the venous system, so requires an oxygenator to facilitate gas exchange. DAP can provide continuous renal blood flow during repairs of the descending thoracic aorta (DTA) or Crawford extent I aneurysms. However, flow is interrupted during the repair of more extensive aneurysms that encompass the renal arteries. Full cardiopulmonary bypass with deep hypothermic circulatory arrest (DHCA) is an alternative strategy that may be used to provide organ protection and create a bloodless surgical field. DHCA is commonly reserved for emergency TAA repairs, or cases in which the application of an aortic cross clamp is not feasible.

**Figure 2.**
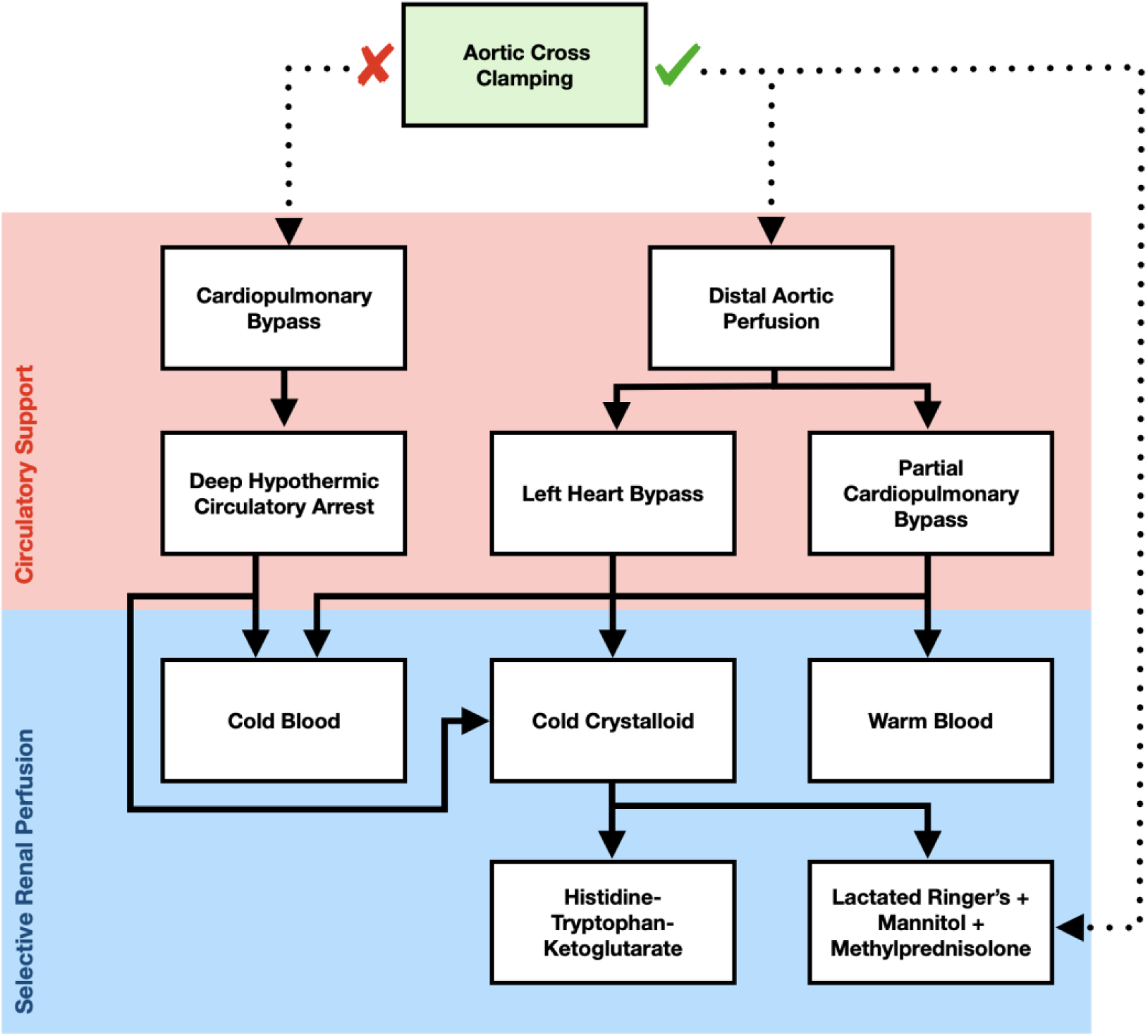
Strategies of circulatory support and selective renal perfusion used during TAA repair. Circulatory support strategies require the use of the heart lung machine for systemic blood flow. Selective renal perfusion involves direct cannulation and perfusion of the renal arteries, and may be used alongside, or independently of, systemic circulatory support.

**Figure 3.**
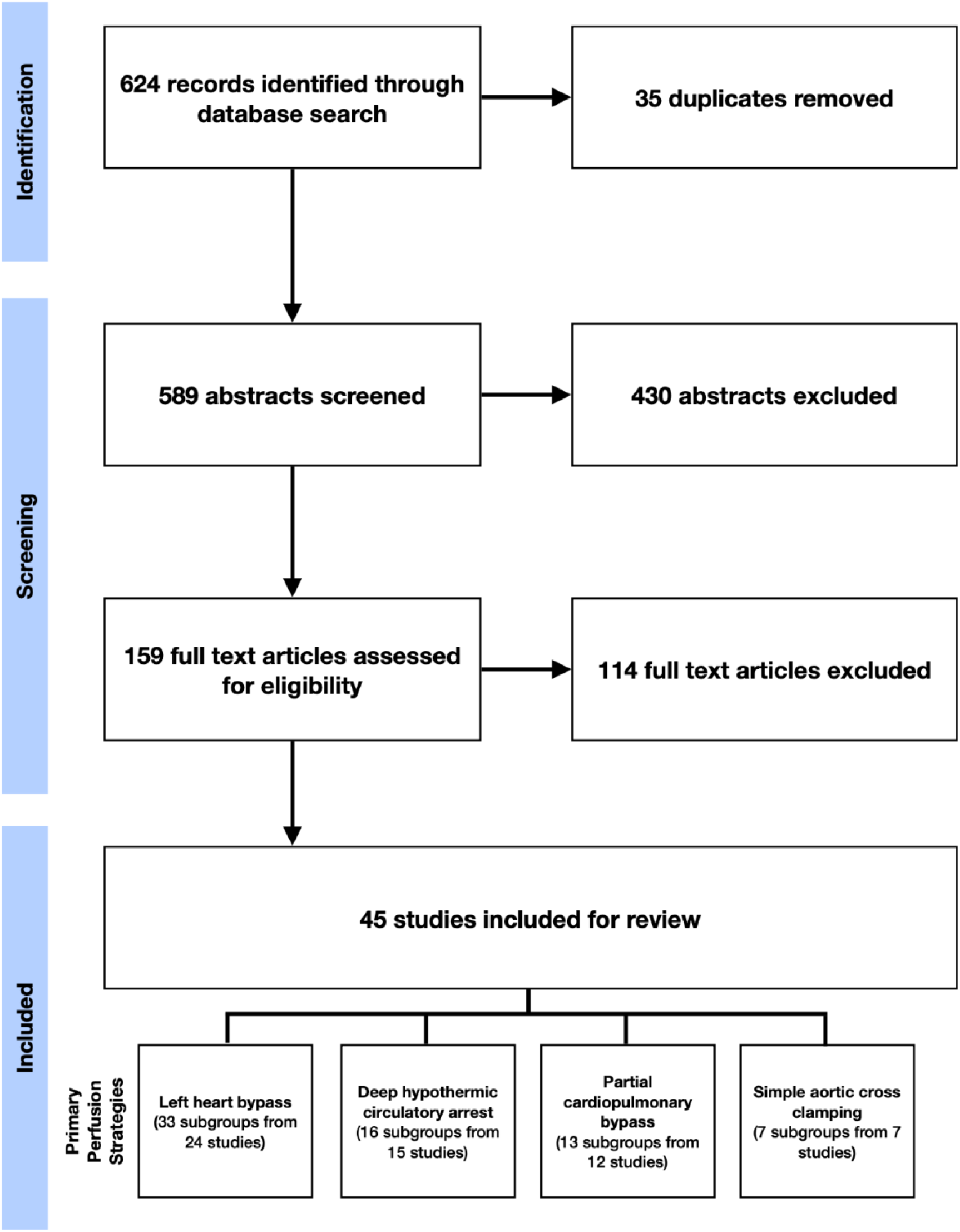
PRISMA flow diagram of the literature search and screening process.

All circulatory support strategies can be augmented with the use of selective renal perfusion (SRP); a technique that involves direct cannulation and perfusion of the renal arteries with warm blood, cold blood or cold crystalloid fluids. ‘Warm’ SRP, delivered at normothermia to mild hypothermia, is intended to maintain renal oxygen delivery; ‘cold’ SRP, routinely cooled to approximately 4°C, aims to reduce renal oxygen consumption^40^. Warm perfusion solutions therefore contain blood to facilitate oxygen delivery, whereas cold solutions can be composed of blood and/or crystalloid fluids. Crystalloid perfusates are commonly enriched with additives designed to increase buffering capacity, maintain cell membrane integrity or reduce inflammation^13^.

A 2010 report from the American College of Cardiology Foundation & American Heart Association recommends that clinicians consider the use of cold blood or crystalloid kidney perfusion during renal artery exposure^39^. Beyond this recommendation, limited evidence-based guidance exists to inform best practice in renal protection. A comprehensive evaluation of intra-operative perfusion techniques is required to establish the efficacy of contemporary renal protection strategies and direct future research and innovation.

This systematic review evaluates published renal and mortality outcomes following TAA repair, to synthesise a narrative discussion on the efficacy of renal protection strategies. Our objectives are:

1. To catalogue the findings of relevant studies published between the years 1995 and 2024.
2. To assess the quality of evidence regarding the use of all featured perfusion techniques.
3. To evaluate the ability of each perfusion technique to ameliorate AKI and operative mortality.
4. To identify important areas of research for future innovation in renal protection.

## Materials and Methods

### Study Protocol and Literature Search

Our protocol was prospectively registered with Prospero (CRD42020166428) and designed in accordance with PRISMA and SWiM recommendations. Searches of PubMed, Web of Science, ClinicalTrials.gov and the EU Clinical Trials Register were conducted to identify relevant English language studies published between 1995 and 2024. Studies were screened against eligibility criteria at the abstract and full-text stages. Included studies were required to feature open surgery on the TAA, an adult patient population, the predominant use of one or more circulatory support strategies, and reporting of post-operative renal outcomes. Studies containing the predominant use of endovascular techniques or type A dissection repairs were excluded. Clinical trials and observational studies were included, whereas single case studies and non-primary publications were excluded. Full literature search terms are provided in table 2.

**Table 2:**
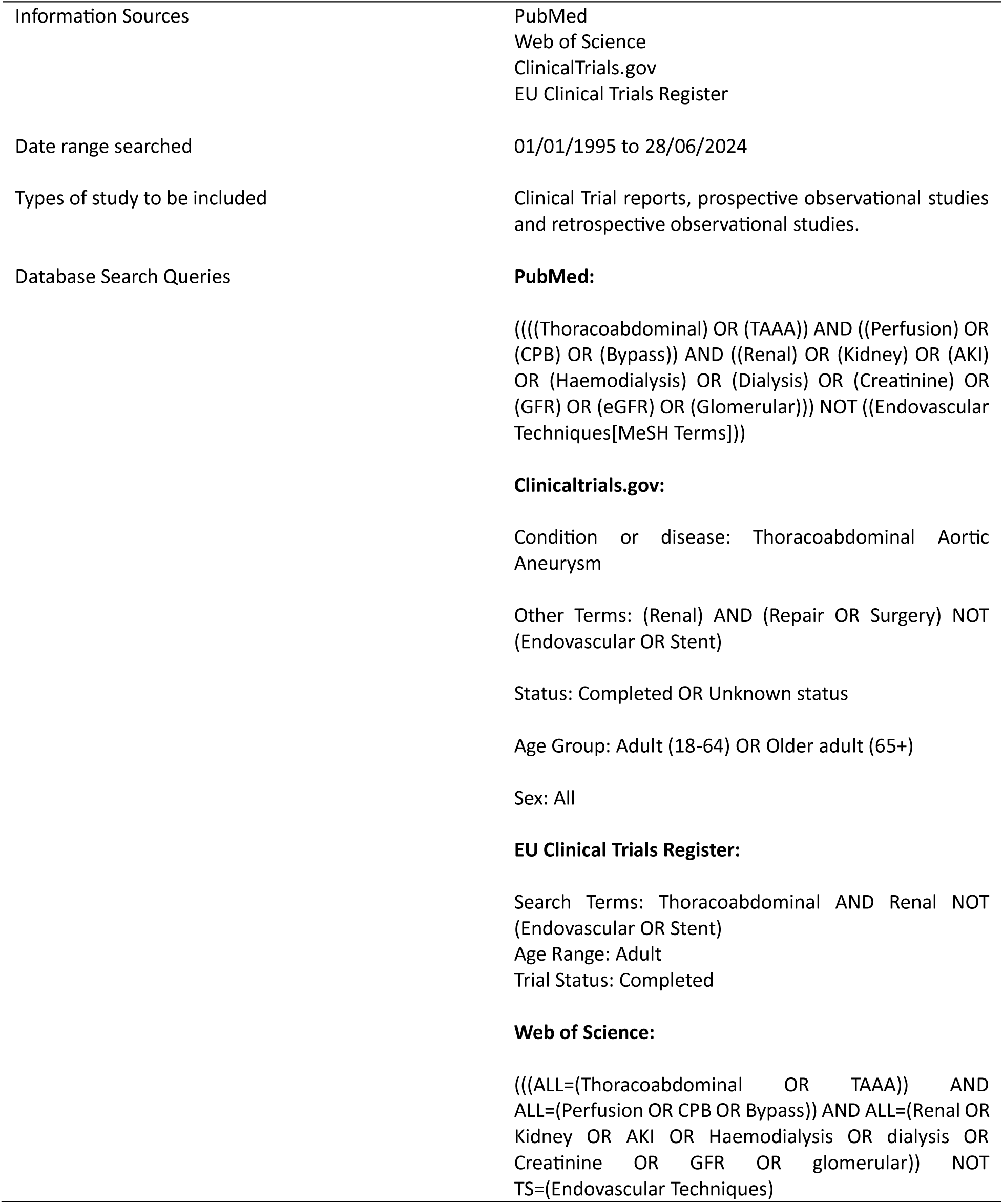
Full Literature Search Terms.

### Data Extraction and Analysis

Data extraction was performed by two reviewers (JB & SS) to ensure consensus in recorded outcomes. Study characteristics, patient demographics, intervention details and clinical outcomes were extracted. Endpoints included the incidence of post-operative RRT, AKI, 30-day mortality and in-hospital mortality. Where appropriate, patient subgroups were identified within studies according to the predominant perfusion strategies used. Data were extracted and presented by patient subgroups, to facilitate the comparison of renal and mortality outcomes by the techniques used. Meta-analysis was not undertaken due to the recognised variability in patient characteristics, operative techniques and reported outcomes present in this area of research.

### Quality and Risk of Bias Assessment

A quality and risk of bias (QROB) assessment was undertaken according to nine risk of bias domains:

▪ Focus on renal protection
▪ Study design and methodology
▪ General representativity/homogeneity of baseline patient characteristics
▪ Risk of participant duplication across multiple studies
▪ Appropriate use of statistical analysis
▪ Certainty of findings, in regard to sample size and magnitude of effect
▪ Standardisation of operative procedures
▪ Detailed account of perfusion techniques
▪ Identified conflicts of interest

Studies were judged to present ‘no concerns’, ‘minor concerns’ or ‘major concerns’ for each QROB criterion. For single cohort observational studies, a judgement of ‘not applicable’ was used in regard to the ‘appropriate use of statistical analysis’. The overall quality of each study was summarised as ‘very high’, ‘high’, ‘moderate’ or ‘low’, based on the number of respective concerns identified (table 3). Following the QROB assessment, a narrative discussion was synthesised, prioritising the findings of the highest quality studies.

**Table 3:** Grading criteria used in quality and risk of bias analysis.

| Table 3: Grading criteria used in quality and risk of bias analysis |  |  |
| --- | --- | --- |
| Overall quality | Number of minor concerns | Number of major or not applicable concerns |
| Very High | ≤1 | 0 |
| High | 2-3 | 0 |
| Moderate | ≤3 | 1 |
| Low | >3 | ≥1 |

## Results

### Study Characteristics & Quality Assessment

45 of 589 identified studies were suitable for inclusion, featuring 3 clinical trials, 7 prospective observational studies (POS) and 35 retrospective observational studies (ROS), representing 6,622 operative cases. A heatmap of participant characteristics is presented in figure 4. 68 study subgroups were identified by the predominant perfusion strategy used. 22 studies featured multiple patient cohorts, enabling the direct comparison of alternate perfusion strategies, whereas 23 studies featured a single patient group.

**Figure 4:**
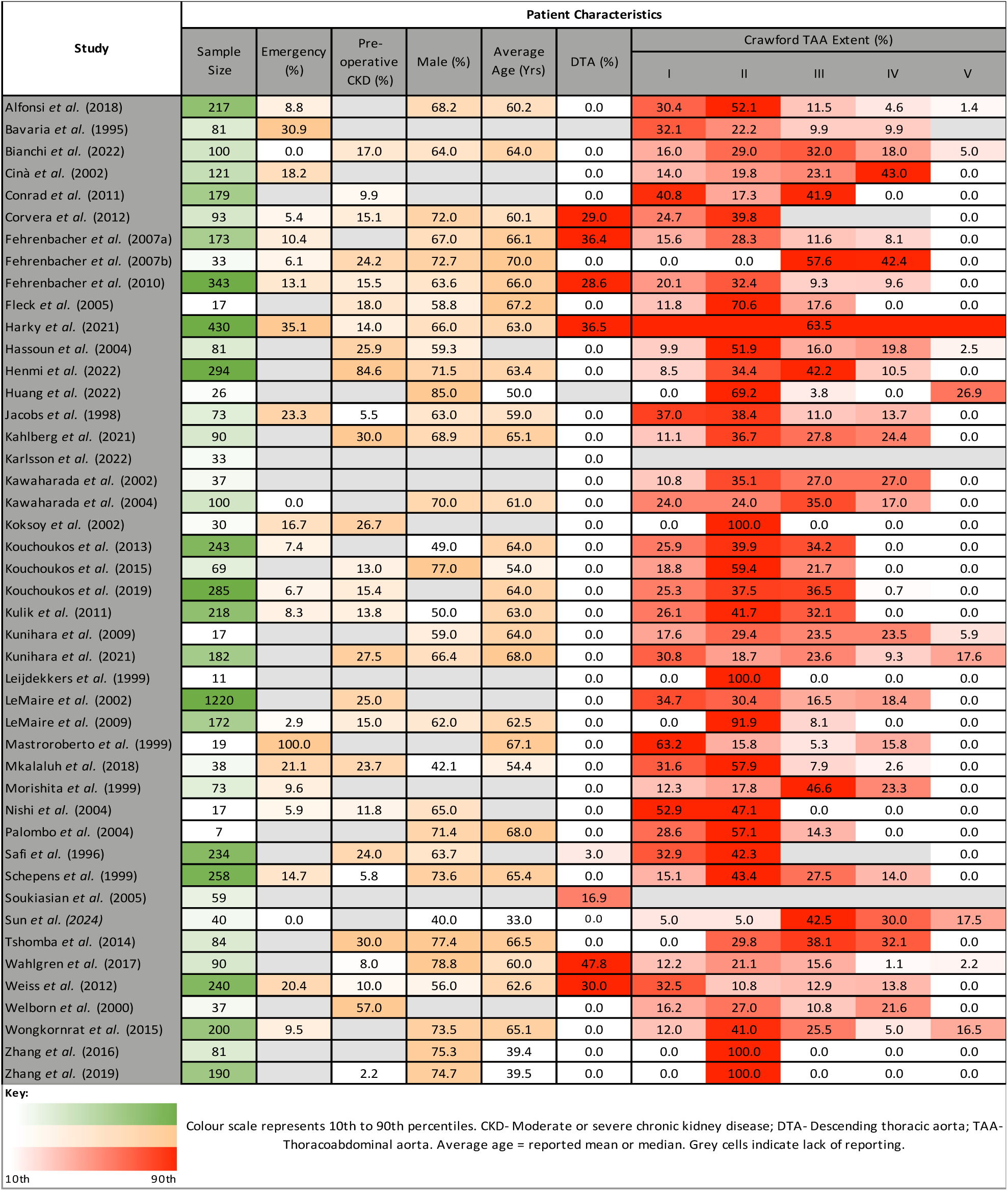
Heatmap of reported patient characteristics from all included studies.

The QROB assessment showed that five studies were of ‘very high quality’, three studies of ‘high quality’, ten studies of ‘moderate quality’ and twenty-seven of ‘low quality’ (figure 5). No significant conflicts of interest were identified.

**Figure 5:**
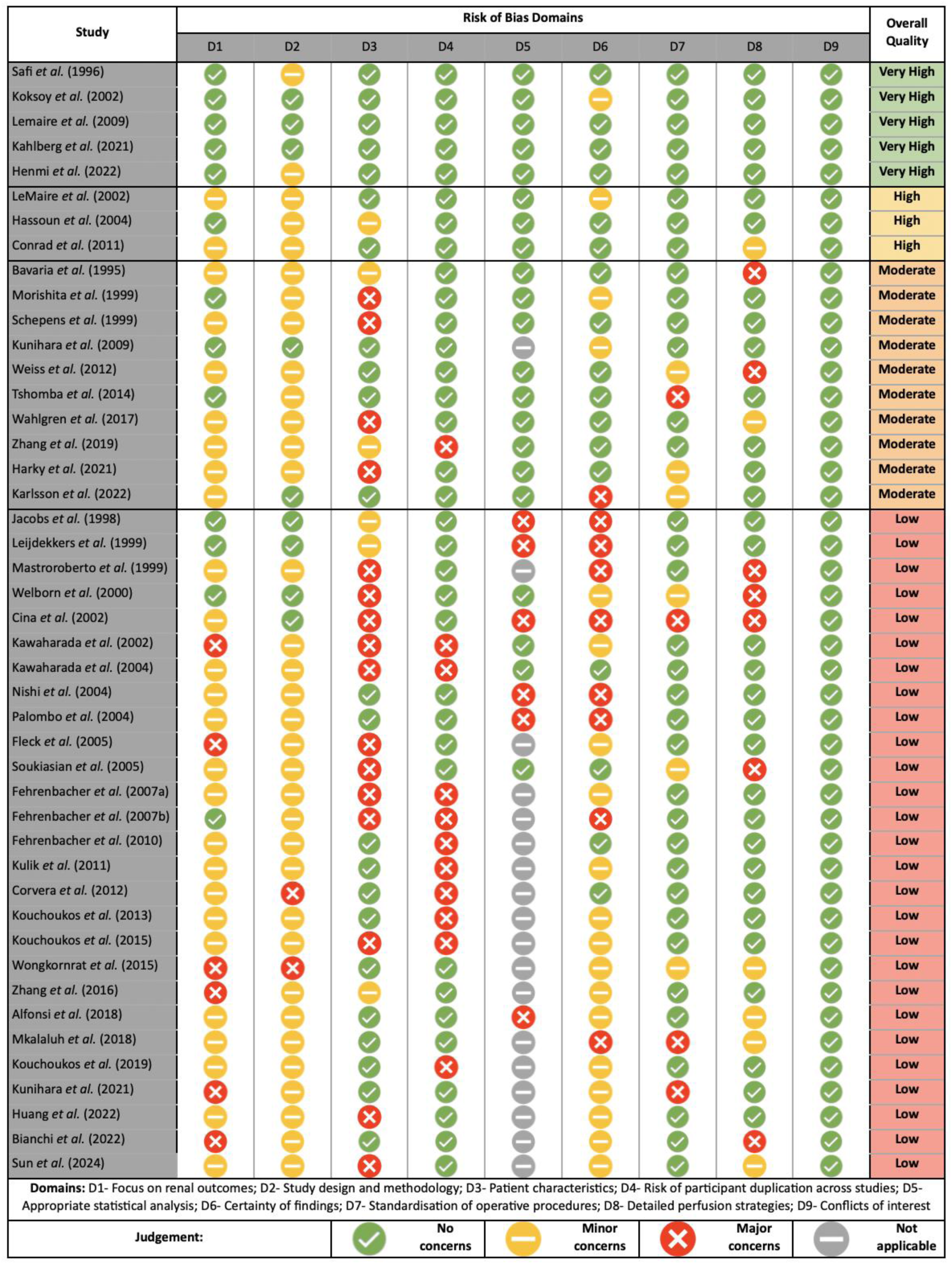
Visualisation of QROB assessment for all included studies.

### Perfusion Techniques

#### Circulatory Support

Three strategies of systemic circulatory support were identified within the 45 included studies (table 4). The exclusive use of LHB was identified in 33 subgroups (24 studies), DHCA in 16 subgroups (15 studies) and pCPB in 13 subgroups (12 studies). 9 subgroups featured mixed perfusion techniques, in which the predominant strategies used were LHB (6 subgroups; 5 studies) and pCPB (3 subgroups; 3 studies). SACC was used exclusively in 7 subgroups (7 studies).

**Table 4.**
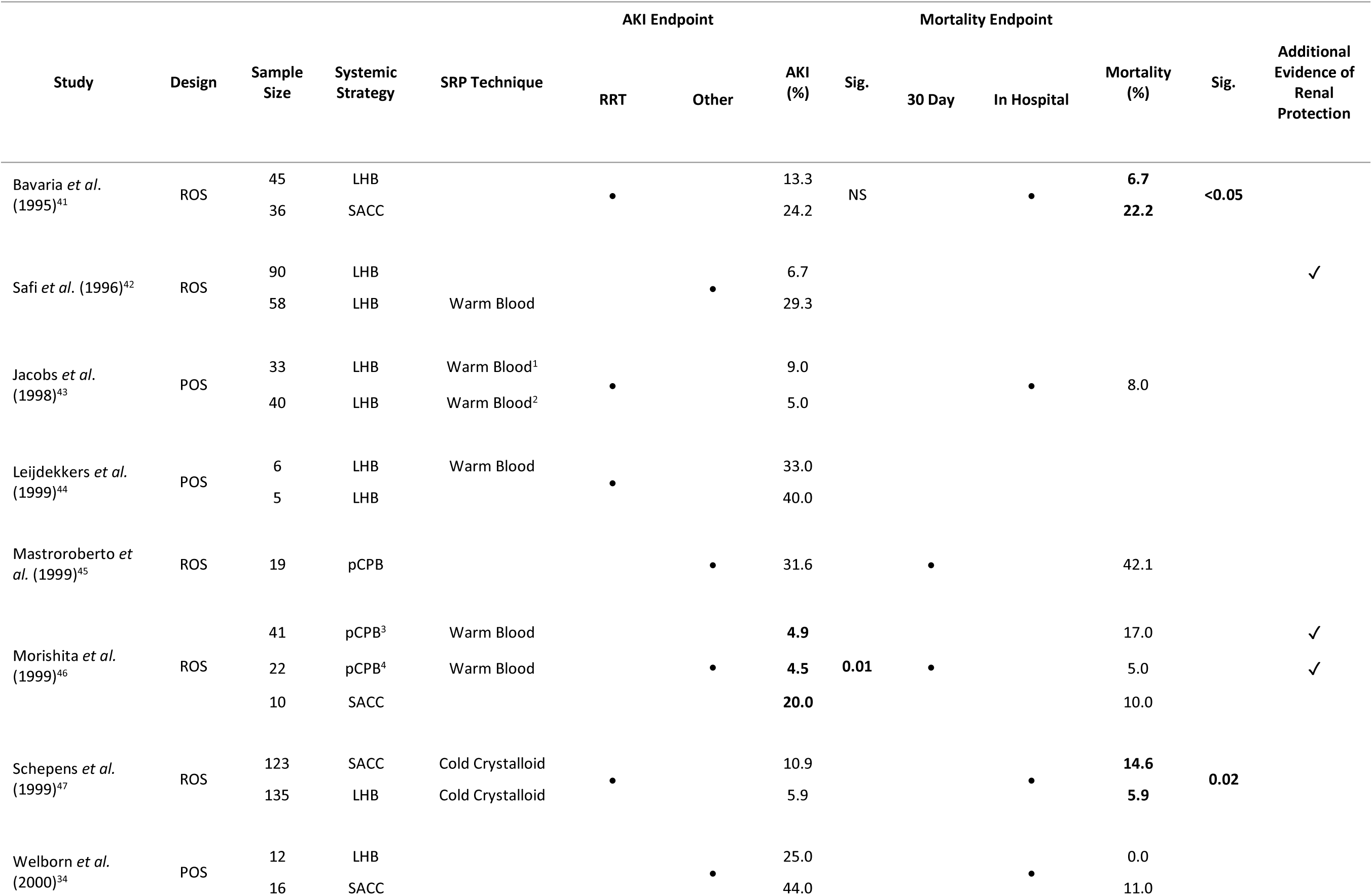
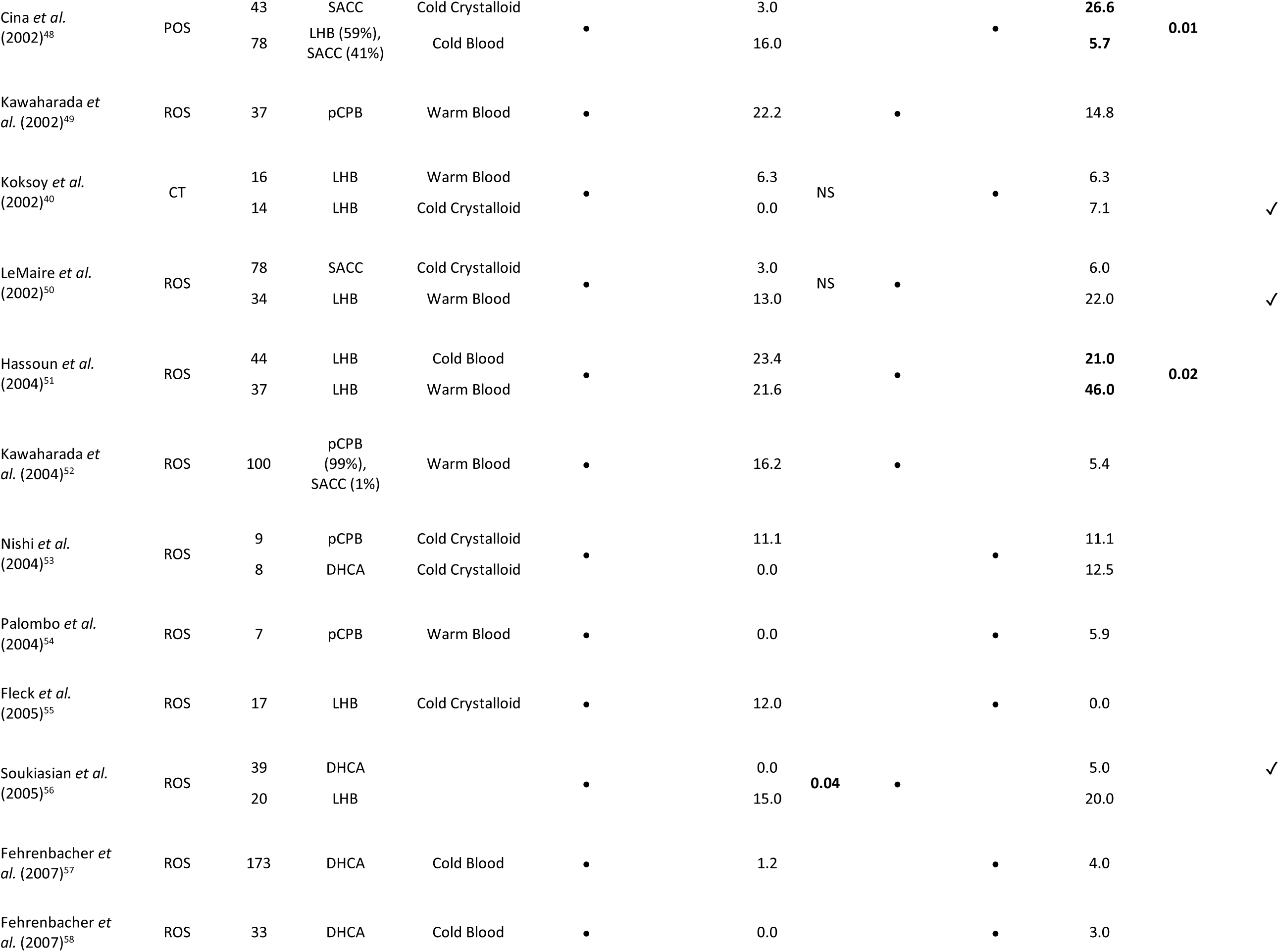
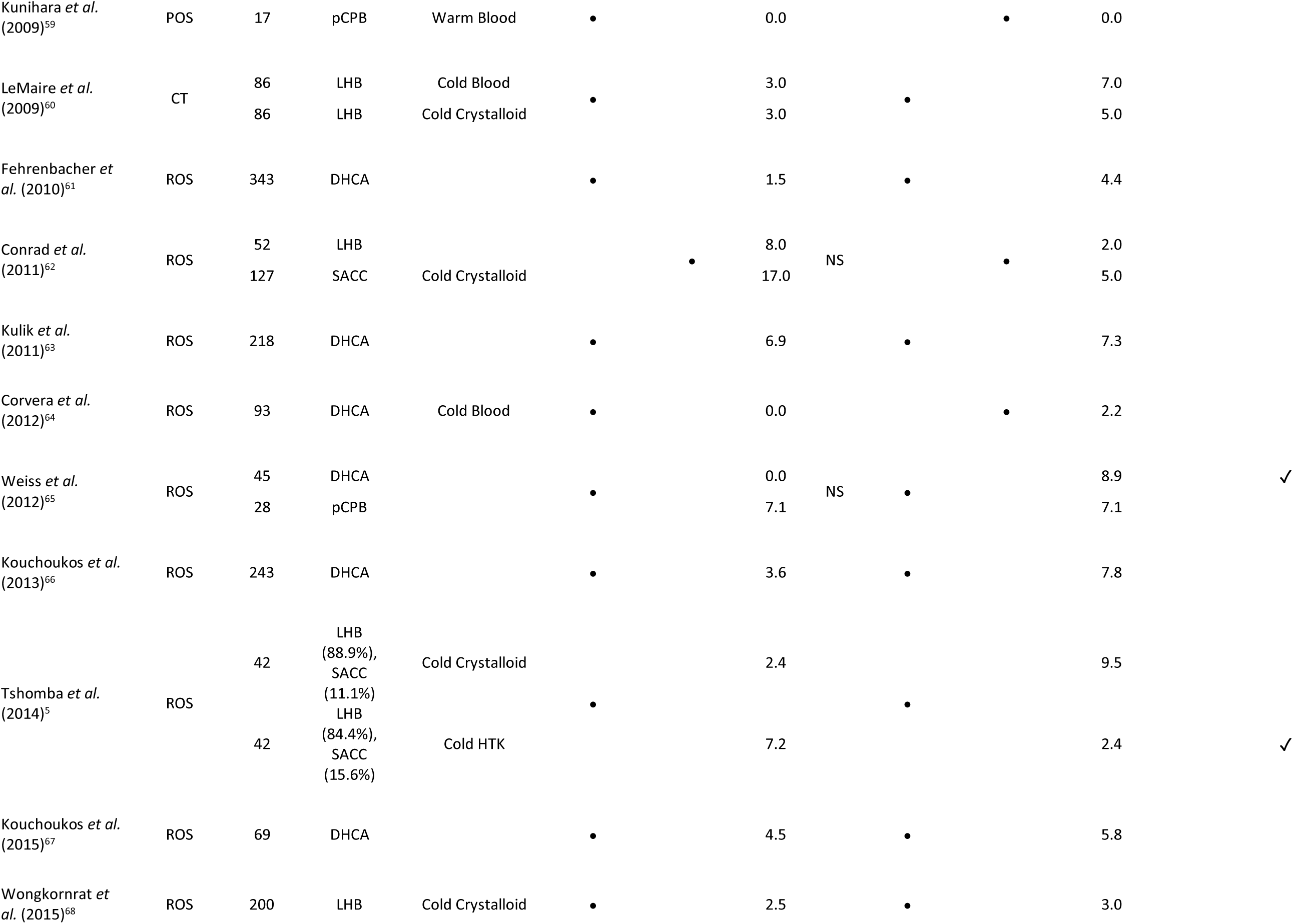
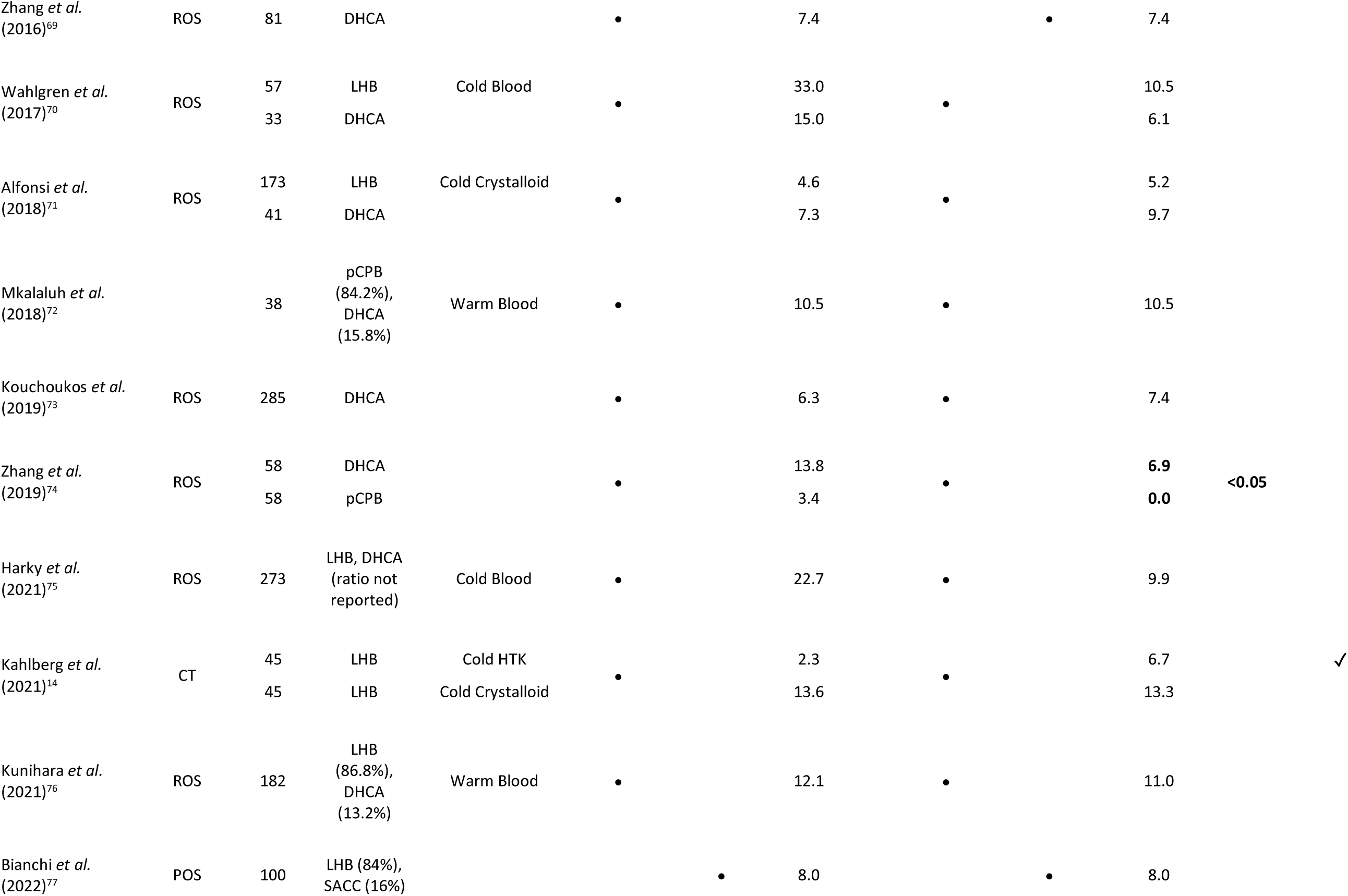
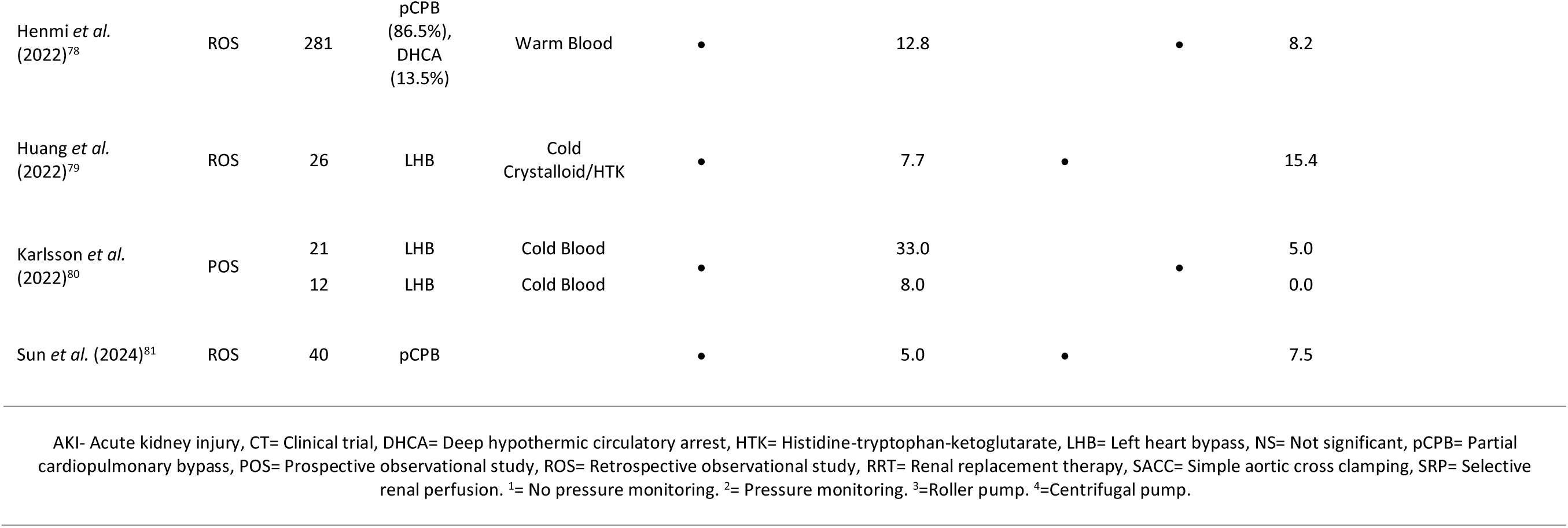
Study Characteristics and Clinical Outcomes.

#### Selective Renal Perfusion

SRP was employed as an adjunct to LHB in 26 subgroups (20 studies), utilising cold crystalloid solutions in 11 subgroups (9 studies), warm blood in 8 subgroups (8 studies) and cold blood in 7 subgroups (7 studies) (table 4). SRP was used alongside pCPB in 9 subgroups (8 studies), including warm blood in 8 subgroups, and cold crystalloid in 1. Five subgroups featured SACC with the adjunctive use of cold crystalloid SRP. Of five subgroups that utilised SRP with DHCA, three used cold blood and one used a cold crystalloid solution. The contents of crystalloid-based renal perfusates are displayed in table 5.

**Table 5.**
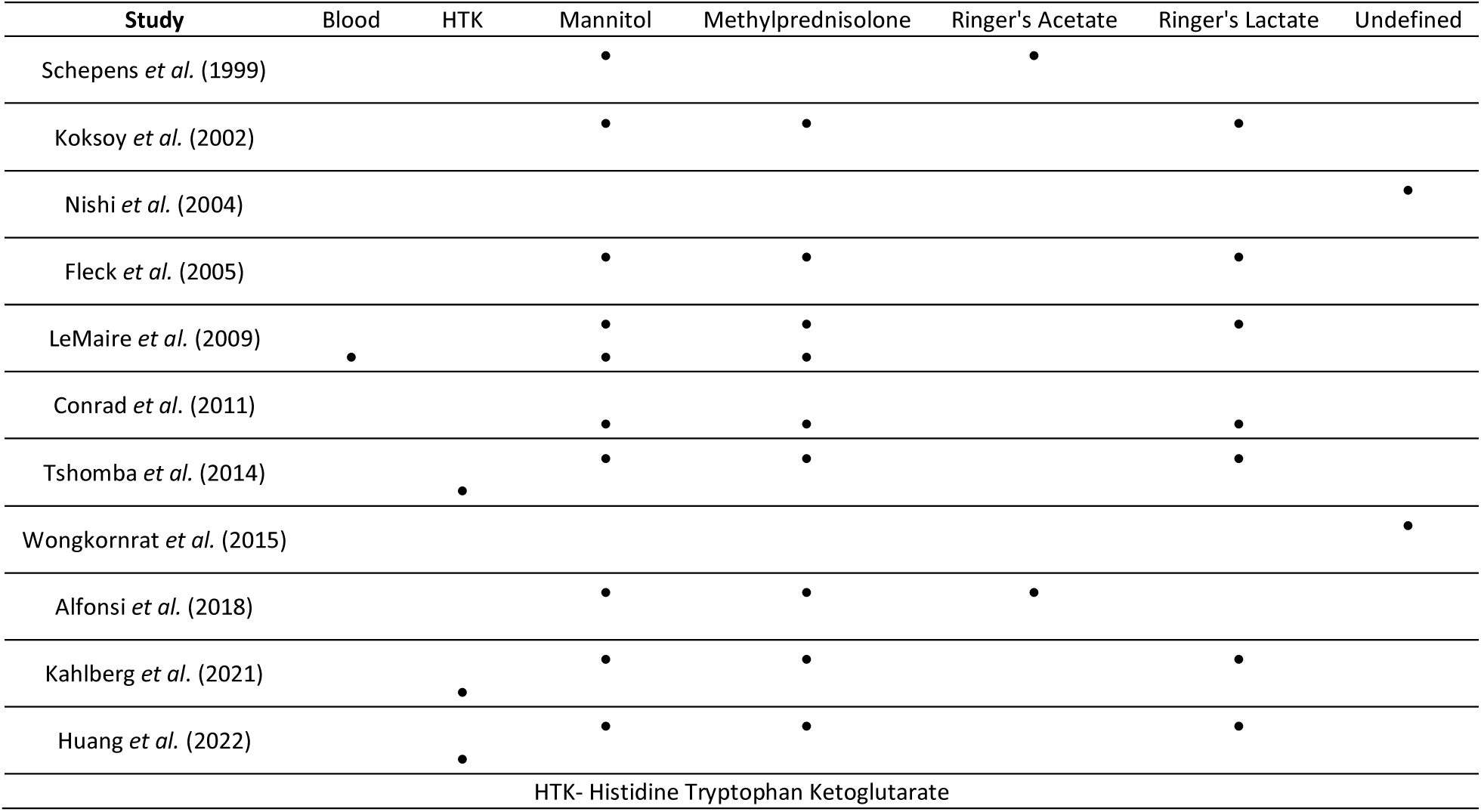
Reported contents of renal perfusates containing crystalloid solutions.

### Outcome Measures

#### Acute Kidney Injury

Several different metrics were identified for the diagnosis of AKI. Post-operative RRT was the most common outcome measure, reported in 39 out of 45 (86.6%) studies, whereas alternative forms of diagnosis were described in 16 (35.5%); these were diverse, and included the use of KDIGO criteria, serum creatinine changes and undefined ‘acute renal failure’. RIFLE or AKIN criteria were not used in any studies. As the incidence of RRT was reported in a majority of studies, it was selected as the primary endpoint for AKI. ‘Other’ AKI endpoints were recorded in the absence of RRT reporting, to facilitate the appropriate comparison of renal injury between cohorts within individual studies. ‘Additional’ evidence of renal protection was reported for studies that included more than one form of renal outcome assessment.

#### Operative Mortality

30-day post-operative mortality was reported in 25 of 45 (55.6%) studies, and ‘in-hospital mortality’ in 18 (40.0%) studies. Both terms are referred to as ‘operative mortality’ in this review. 2 (4.4%) studies did not report mortality outcomes.

### Outcomes by Perfusion Strategy

Renal and mortality endpoints associated with each study subgroup are presented in table 4. Where a single circulatory support technique was used for over 75% of patients per study subgroup, renal and mortality endpoints and displayed in figures 6 & 7 by the predominant technique used.

**Figure 6.**
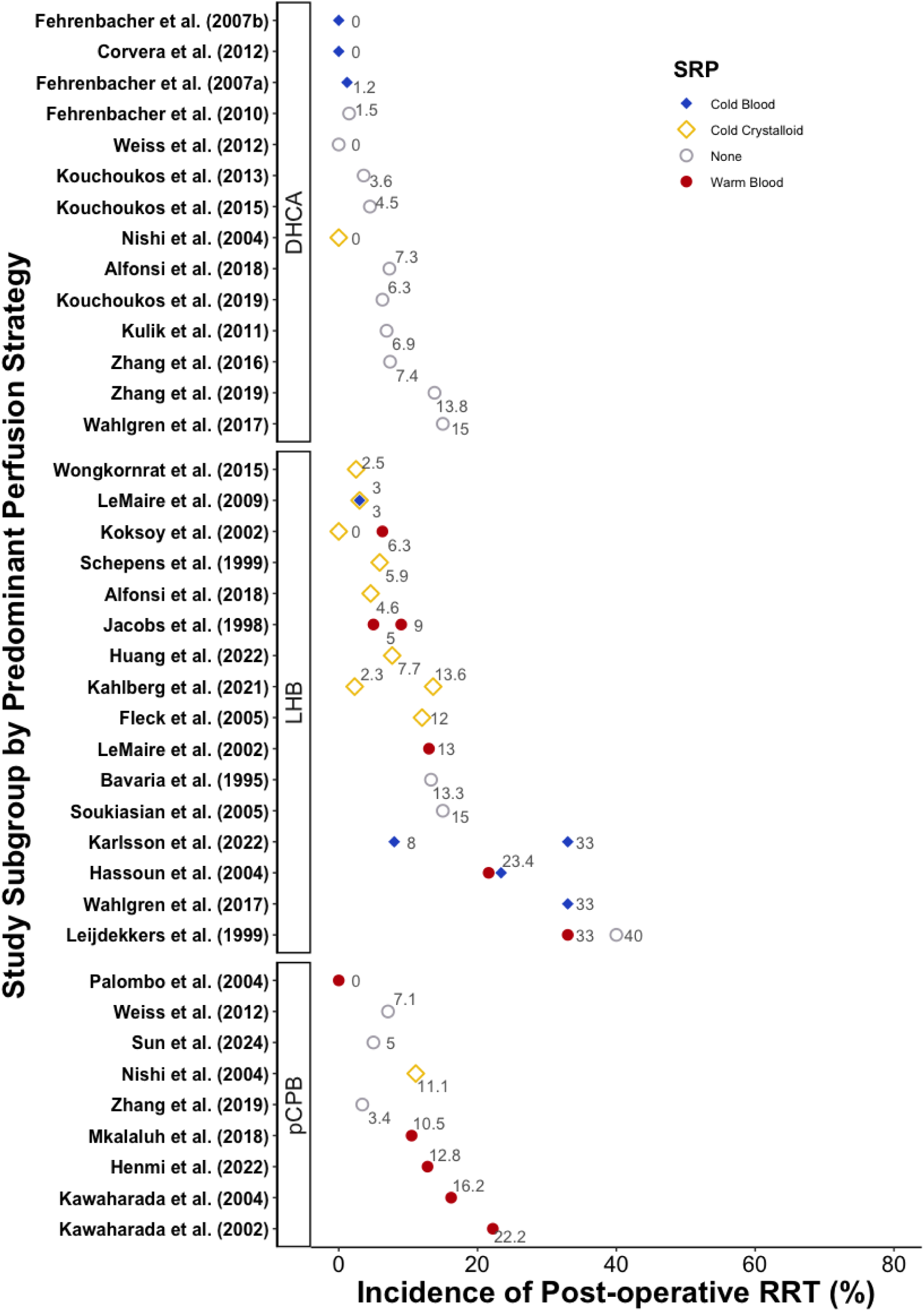
The reported incidence of post-operative renal replacement therapy by study subgroup, according to strategies of circulatory support and selective renal perfusion. DHCA-cardiopulmonary bypass with deep hypothermic circulatory arrest; LHB-left heart bypass; pCPB-partial cardiopulmonary bypass; SRP-selective renal perfusion.

**Figure 7.**
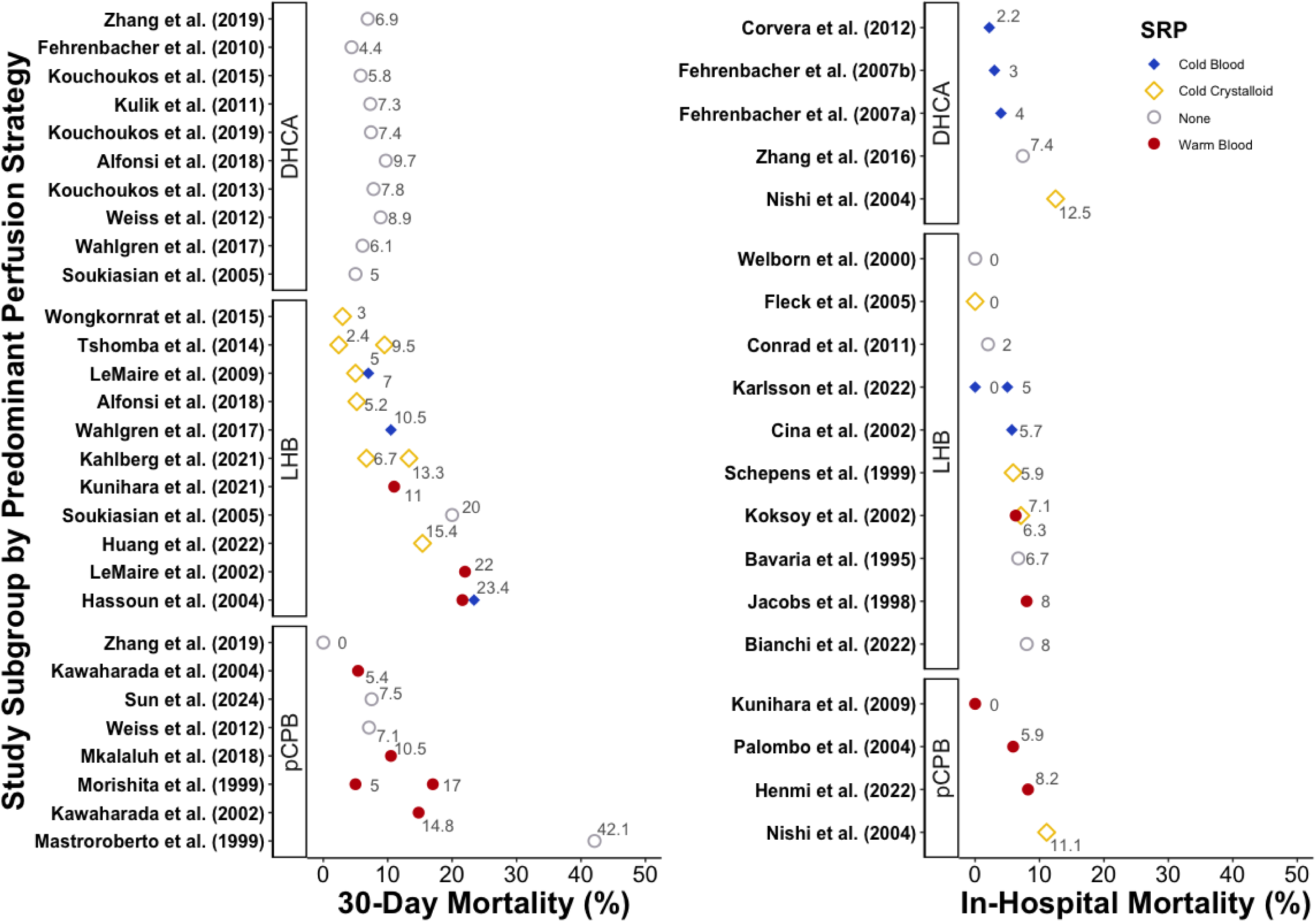
The reported incidences of 30-day and in-hospital mortality by study subgroup, according to strategies of circulatory support and selective renal perfusion. DHCA-cardiopulmonary bypass with deep hypothermic circulatory arrest; LHB-left heart bypass; pCPB-partial cardiopulmonary bypass; SRP-selective renal perfusion.

#### Distal Aortic Perfusion: Left Heart Bypass

The use of LHB *without* adjunctive SRP was associated with a 13.3-40.0% incidence of post-operative RRT, and a 0-20.0% risk of operative mortality. When used *with* adjunctive SRP, LHB was associated with incidences of RRT ranging from 5.0% to 33.0% with the use of normothermic blood, 3.0% to 33.0% with cold blood, and 0% to 13.6% with cold crystalloid solutions. Operative mortality ranged from 6.3% to 22.0% with the use of warm blood SRP, 5.7% to 10.5% with cold blood, and 0% to 15.4% with cold crystalloid solutions.

Four studies demonstrated that LHB can be used to achieve outcomes superior to SACC. Bavaria and colleagues showed that LHB was associated with a reduced incidence of in-hospital mortality (6.7% vs 22.2%, p<0.05)^41^. Schepens and colleagues found that the odds of requiring post-operative RRT were 92% lower with LHB than SACC (O.R. 0.08, 95%C.I. 0.01-0.6, p=0.02)^47^. LHB was also associated with a lower incidence of operative mortality (5.9% vs 14.6%, p<0.02)^47^. LeMaire and colleagues demonstrated that LHB could improve rates of AKI compared to SACC (9% vs 32%, p<0.0005), despite the adjunctive use of cold crystalloid SRP alongside SACC. This protection was also associated with non-significant reductions in RRT (3% vs 13%, p=0.167) and 30-day mortality (6% vs 22%)^50^. Similarly, Conrad and colleagues (2011) utilised propensity-matching to show that AKI (8% vs 17%, p=0.10) and in-hospital mortality (2% vs 5%, p=0.38) were lower with LHB than SACC with cold crystalloid SRP. However, their findings did not reach statistical significance^62^. Together, these findings support the use of DAP for the amelioration of renal IRI.

The use of SRP has been employed alongside LHB in several studies, to varying degrees of success. Warm blood SRP is generally associated with inferior outcomes, whereas selective renal cooling has been shown to provide effective kidney protection. In 1996, Safi and colleagues compared LHB with warm blood SRP to a non-adjunctive strategy of LHB, and identified warm blood SRP as a risk factor for AKI, associated with a four-fold increase in its incidence^42^. The mean collective blood flow was 250ml/min, shared between the renal and visceral arteries. Similarly, Koksoy and colleagues compared warm blood SRP to cold crystalloid SRP during LHB to find that warm blood SRP was associated with a three-fold rise in AKI (63% vs 21%, p=0.03)^40^. Hassoun and colleagues (2004) compared the use of warm and cold blood SRP during LHB, discovering that cold blood SRP was associated with a significantly lower incidence of operative mortality (27% vs 56%, p<0.02), despite no significant differences in recovery from renal dysfunction during hospitalisation (27% warm vs 36% cold, p=0.37)^51^. In a 2009 clinical trial, LeMaire and colleagues compared intermittent boluses of cold blood or crystalloid solution with additives mannitol and methylprednisolone during LHB. They found that the rate of RRT was identical in both cohorts (3.0% vs 3.0%, p=1.0), and there was no difference in the incidence of 30-day mortality (7.0% blood vs 5.0% crystalloid, p=0.7)^60^. Thus, there is strong evidence that renal cooling with blood or crystalloid solutions provides equivalent protection from IRI.

Furthermore, the contents of crystalloid perfusates can affect kidney protection. In a 2021 clinical trial, Kahlberg and colleagues compared the use of HTK solution to a Ringer’s-based crystalloid perfusate, and demonstrated a lower incidence of KDIGO AKI with HTK solution (48.9% vs 75.6%, p=0.02), though rates of RRT were lower but not significantly different (2.2% vs 13.3%, p=0.11)^14^. Additionally, no significant difference in 30-day mortality was identified (6.7% HTK vs 13.3% Ringer’s, p=0.23)^14^.

#### Distal Aortic Perfusion: Partial Cardiopulmonary Bypass

The use of pCPB *without* SRP was associated with a 3.4-7.1% incidence of post-operative RRT and a 0-7.1% risk of operative mortality. pCPB with adjunctive warm blood SRP was associated with a 0-22.2% incidence of RRT. One study that utilised cold crystalloid SRP was associated with 11.1% incidences of RRT and in-hospital mortality.

Like LHB, the use of pCPB can provide superior renal protection than SACC. Morishita and colleagues demonstrated that pCPB with adjunctive warm blood SRP could be used to significantly reduce the risk of RRT, irrespective of the type of blood pump used for circulatory support (4.5% centrifugal pump, 4.9% roller pump, 20% SACC, p=0.01). pCPB was also associated with higher intraoperative urine production (2.9ml/hr roller SRP, 2.8ml/hr centrifugal SRP, 0.2ml/hr SACC) and lower post-operative serum creatinine concentrations (p<0.0001) than SACC^46^. With a small cohort of patients, Kunihara and colleagues (2009) achieved no incidences of RRT or operative mortality with pCPB and warm blood SRP^59^. More recently, Henmi and colleagues (2022) analysed the results from 281 cases to show that 150-200ml/min of warm blood SRP per renal artery was associated with a 12.8% incidence of RRT, a 63.3% incidence of KDIGO AKI and an 8.2% risk of in-hospital mortality^78^. Notably, this study included a high (84.6%) incidence of pre-operative renal dysfunction.

A propensity-matched comparison study from Weiss and colleagues (2012) found no significant difference in the rate of RRT following pCPB or DHCA, though DHCA was associated with a lower incidence of AKI (22.2% vs 46.4%, p=0.03)^65^. Notably, a higher proportion of DHCA patients received DTA repair (22% vs 42%, p=0.04), which is generally associated with reduced renal ischaemic times than TAA repair. Similarly, a propensity-matched study by Zhang and colleagues (2019) compared pCPB to DHCA and showed that whilst pCPB was associated with lower risks of RRT (3.4% vs 13.8%, p=0.047) and 30-day mortality (0% vs 6.9%, p<0.05), there was no significant difference in the rate of new-onset renal insufficiency (13.8% vs 24.1%, p=0.126)^74^.

#### Deep Hypothermic Circulatory Arrest

The use of DHCA is associated with low incidences of RRT (0-15.0%) and operative mortality (2.2-12.5%). Soukiasian and colleagues (2005) compared DHCA to LHB and demonstrated that a reduced incidence of AKI, defined by a 1.5-fold increase in serum creatinine concentration, was associated with DHCA (0% vs 15%, p=0.04)^56^. Wahlgren and colleagues (2017) compared the use of DHCA with LHB and cold blood SRP, but found no significant differences in rates of AKI or 30-day mortality^70^. Consequently, there is evidence to suggest that DHCA can provide a high standard of renal protection. However, despite several studies of reasonable sample size, no study on DHCA was judged to present high-quality evidence (figure 5). Further prospective research into the effect of DHCA on kidney protection is therefore warranted, to facilitate the appropriate review of this technique.

## Discussion

Surgical repair of the TAA is a life-saving procedure complicated by high risks of AKI and operative mortality. Addressing these issues requires the use of effective intraoperative kidney protection strategies. This systematic review assesses the findings of 45 published studies to identify the incidences of AKI and operative mortality by different perfusion techniques. Furthermore, through quality and risk of bias analysis, we evaluate the strength of evidence supporting the use of each technique.

The findings of this review should be interpreted with caution due to the limited number of studies deemed to present high or very-high quality evidence. The majority of studies were judged to be low quality, with 35 studies being of retrospective design and 23 featuring a single patient cohort. Just three randomised clinical trials have been conducted in this field, and with the evident heterogeneity in patient characteristics and operative techniques, we were limited in our ability to directly compare the efficacy of alternative perfusion strategies. Furthermore, the 29-year time period within which the included studies were published was host to an evolution in the criteria used to define AKI. Study endpoints therefore demonstrate a considerable level of variability.

Despite these constraints, several key themes emerged within our analysis of moderate to very-high quality studies:

1. Distal aortic perfusion provides effective kidney protection
2. Adjunctive renal cooling can enhance kidney protection during extensive TAA repairs
3. Warm selective renal perfusion provides inferior kidney protection
4. Deep hypothermic circulatory arrest provides promising kidney protection, but requires further clinical investigation

### 1. DAP provides effective kidney protection

Normothermic to mild hypothermic perfusion of the distal aorta provides superior kidney protection than SACC. LHB and pCPB can both provide DAP, perfuse the renal arteries during suprarenal aortic clamping and reduce exposure of the kidneys to IRI. During repairs of the DTA or extent I TAA, DAP can maintain kidney perfusion for the duration of surgery, eliminating renal IRI, as clamping of the infrarenal aorta is not required. During the repair of Crawford extent II-IV aneurysms, however, infra-renal clamping is necessary, meaning that renal IRI is unlikely to be avoided with an isolated strategy of DAP. The combined use of sequential aortic clamping may help to reduce renal ischaemic time^21^.

### 2. Adjunctive Renal Cooling can enhance Kidney Protection during extensive TAA repairs

When DAP cannot prevent kidney ischaemia, the renal arteries may be directly cannulated to facilitate perfusion of the kidneys with blood or crystalloid solutions. SRP may feature normothermic blood to support kidney oxygenation, or cold perfusates to enable hypothermic renal protection. Hypothermia is an established cryoprotection strategy, used during cardiopulmonary bypass to reduce cellular metabolism and associated oxygen demands^82^.

Our findings demonstrate that cold blood and crystalloid solutions - generally composed of lactated Ringer’s solution, mannitol and methylprednisolone - can deliver equivalent protection from AKI and operative mortality^60^. Moreover, the use of HTK solution can further improve protection, and is linked to a low incidence of post-operative RRT^14^. However, such strategies should be used with care, as extensive cold crystalloid perfusion is associated with risks of systemic hypothermia and haemodilution, and may expose patients to an increased risk of cardiac arrest during surgery. Furthermore, this risk may be augmented with HTK solution, which is cardioplegic in nature, and could cause systemic hyponatraemia, arrhythmias or cardiac arrest if managed inappropriately. Finally, cold fluids also expose the kidneys to a sudden warm, highly oxygenated environment on reperfusion, which is theorised to exacerbate renal injury^83^. Whilst cold SRP can therefore be used to improve renal protection and reduce the incidence of AKI following surgery, it should be used with care. There is currently no professional guidance on the management of HTK solution for kidney protection, and further research into its safe and effective use is advised.

### 3. Warm Blood Selective Renal Perfusion provides inferior kidney protection

The theoretical advantages of normothermic blood SRP include functional protection of the kidneys and a reduced risk of cardiac arrest. However, our findings demonstrate that its use is associated with inferior renal protection and increased mortality. Several studies conducted out of Houston, Texas collectively illustrate that warm blood SRP used during LHB provides poor renal protection^40, 42, 51^. Similarly, whereas warm blood SRP used alongside pCPB may provide superior kidney protection than SACC^46^, a majority of patients still develop KDIGO AKI^78^.

It is likely that historic applications of warm SRP were performed sub-optimally. The kidneys are metabolically active organs that typically receive a combined blood flow of 1200ml/min in normal physiology^84^. Within the studies of this review, the maximum reported flow provided to the kidneys was 300ml/min per artery^40^. Furthermore, studies typically employed the use of a branched catheter system to enable simultaneous perfusion of the superior mesenteric and celiac arteries, which could lead to preferential flow away from the kidneys due to high relative renal resistance or narrow renal perfusion cannulae. It is feasible that flow rates used in these studies did not provide a sufficient delivery of oxygen to meet the metabolic demands of the kidneys at normothermia. Within the related discipline of *ex vivo* renal perfusion for transplantation, Weissenbacher and colleagues have achieved functional normothermic perfusion of human kidneys for 24 hours at a mean flow of 364ml/min per kidney, producing a mean urine output of 27.4ml/hr^85^. This represents a higher flow than that used in any of the studies included in this review. Whilst the clinical impact of normothermic perfusion cannot be determined from the results of Weissenbacher’s study, the findings demonstrate that adequate blood flow can sustain functional protection. Furthermore, the incorporation of pulsatile flow may offer an additional advantage to kidney perfusion, but also requires further research and development prior to clinical implementation^86, 87^.

In TAA surgery, SRP cannot be continued for the entire duration of surgery, and must be stopped during reattachment of the renal arteries. During this time period, a warm kidney will be exposed to a greater degree of IRI than a kidney protected by a cold preservation solution. A combined strategy of functional, normothermic SRP followed by a bolus of hypothermic perfusate may provide effective intraoperative kidney protection. Without further pre-clinical optimisation, the results of this review do not support the use of warm SRP for kidney protection.

### 4. DHCA requires further clinical investigation

The establishment of deep hypothermia can reduce global metabolic demand and enable arrest of the systemic circulation^88^. Several studies demonstrate incidences of AKI comparable to the use of LHB with cold crystalloid SRP, suggesting that DHCA is an encouraging technique for kidney protection. However, the evidence for the use of DHCA is restricted to retrospective observational studies from a limited number of surgical centres. Furthermore, additional clinical outcomes and implications should be considered in the evaluation of novel care practices, and the use of DHCA is reportedly linked to higher rates of post-operative bleeding, stroke, low cardiac output syndrome and prolonged ventilator support than normothermic DAP^7, 89^. Moreover, within this series of reviewed papers, DHCA was the only circulatory support strategy not capable of achieving a 0% rate of operative mortality. Prospective research into the use of DHCA is recommended in order to determine the full implications of its use.

### Intravascular Haemolysis and Myoglobinaemia

Evidently, the use of any perfusion strategy is limited in its ability to protect patients from AKI. Whilst clamp-induced renal IRI is understood to be the primary cause of AKI, additional risk factors are known to contribute to its onset, and should be considered in any comprehensive package of renal protection. Importantly, high concentrations of circulating hemeproteins, such as haemoglobin and myoglobin, are associated with impaired post-operative renal function.

In 2022, Kiser and colleagues identified the intensive use of cell salvage during LHB as a risk factor for AKI and operative mortality^90^. The common strategy of reinfusing unwashed, salvaged blood into the patient circulation is a likely cause of significant intravascular haemolysis. Cardiotomy suction causes red blood cell damage and the release of free haemoglobin into plasma. Elevated free haemoglobin concentration has been identified as a risk factor for AKI following cardiopulmonary bypass for TAA repair, and Extra-Corporeal Membrane Oxygenation (ECMO)^4, 30^. Plasma free haemoglobin can scavenge nitric oxide present within the renal vascular endothelium, inhibit vasodilation and augment renal ischaemia. Furthermore, heme, a product of haemoglobin oxidation, has been shown to cause direct proximal tubular cell toxicity, and can trigger an inflammatory cascade leading to cell damage^3, 35^. Haemolysis may therefore contribute to AKI via renal endothelial dysfunction, cellular toxicity and amplification of the inflammatory response to surgery. Similarly, myoglobinaemia, caused by ischaemia of the femoral arteries, is known to contribute to AKI^91, 92^. The integration of perfusion techniques that minimise blood damage and peripheral ischaemia are therefore key to providing a holistic system of care that reduces renal hypoxia and toxicity.

### Study Limitations

Whilst this review provides a comprehensive analysis of published studies on renal protection during TAA surgery, its findings should be interpreted within the context of its inherent limitations. For example, results are limited to a crude analysis of renal and mortality outcomes subject to variations in patient characteristics, variability in the quality of reported details, and inconsistent AKI diagnosis criteria. Furthermore, no other morbidities, nor long-term mortality outcomes were analysed, but should be incorporated in any rigorous system of clinical decision-making. Similarly, we do not analyse certain surgical practices that may influence renal outcomes, such as patch vs Coselli graft repairs or the use of sequential clamping, renal stenting or endarterectomy. Nor do we include an analysis of warm renal ischaemic times, due to a general scarcity in the reporting of this information. Finally, whilst clinical outcomes were independently extracted and judged for consensus by two reviewers, the screening and quality assessment stages were performed by a single reviewer who adhered to a pre-determined set of screening questions (JB).

## Conclusion

AKI is a common concern following TAA surgery and is linked to an increased risk of operative mortality. Whilst the pathophysiology of AKI is multifactorial, it is commonly stimulated by clamp-induced renal IRI, which may be compounded by intravascular haemolysis and myoglobinaemia. The evidence from our analysis supports the use of distal aortic perfusion for extent I TAA repairs, and adjunctive cold SRP with HTK solution for extent II, III and IV TAA repairs. Normothermic SRP cannot be advocated on the basis of this review, but warrants pre-clinical investigation. Cardiopulmonary bypass with DHCA is an alternative strategy that may be capable of providing an excellent kidney protection, however, its use is supported by low-quality evidence and may be associated with additional adverse outcomes. Further, high-quality comparative research is necessary to establish its efficacy and limitations in relation to DAP and SRP.

## Data Availability

All data produced in the present study are available upon reasonable request to the authors.

## Non-standard abbreviations and acronyms

AKI: Acute kidney injury
AKIN: Acute kidney injury network
ATN: Acute tubular necrosis
ATP: Adenosine Triphosphate
CPB: Cardiopulmonary bypass
DAP: Distal aortic perfusion
DHCA: Deep hypothermic circulatory arrest
ECC: Extracorporeal circulation
EU: European Union
HLM: Heart lung machine
IRI: Ischaemia-reperfusion injury
KDIGO: Kidney disease improving global outcomes
LHB: Left heart bypass
pCPB: Partial cardiopulmonary bypass
POS: Prospective observational study
PRISMA: Preferred reporting items for systematic reviews and meta-analyses
QOL: Quality of Life
RCT: Randomised controlled trial
RIFLE: Risk, Injury, Failure, Loss, End Stage Renal Disease
ROB: Risk of bias
ROS: Retrospective observational study
RRT: Renal replacement therapies
SACC: Simple aortic cross clamping
SRP: Selective renal perfusion
SWiM: Synthesis without meta-analysis
TAA: Thoracoabdominal aorta

## Acknowledgements

None

